# Preliminary estimating the reproduction number of the coronavirus disease (COVID-19) outbreak in Republic of Korea and Italy by 5 March 2020

**DOI:** 10.1101/2020.03.02.20030312

**Authors:** Zian Zhuang, Shi Zhao, Qianying Lin, Peihua Cao, Yijun Lou, Lin Yang, Shu Yang, Daihai He, Li Xiao

## Abstract

The novel coronavirus disease 2019 (COVID-19) outbreak and Italy has caused 6088 cases and 41 deaths in Republic of Korea and 3144 cases and 107 death in Italy by 5 March 2020. We modeled the transmission process in Republic of Korea and Italy with a stochastic model and estimated the basic reproduction number *R*_0_ as 2.6 (95% CI: 2.3-2.9) or 3.2 (95% CI: 2.9-3.5) in Republic of Korea, under the assumption that the exponential growth starting on 31 January or 5 February 2020, and 2.6 (95% CI: 2.3-2.9) or 3.3 (95% CI: 3.0-3.6) in Italy, under the assumption that the exponential growth starting on 5 February or 10 February 2020. Estimates of dispersion term (k) were 10 (95% CI: 5-56) or 22 (95% CI: 8-61) in Republic of Korea, and 13 (95% CI: 5-61) or 37 (95% CI: 13-61) in Italy, and all of which imply few super-spreading events.

## 1. Introduction

The coronavirus disease 2019 (COVID-19) first emerged in Wuhan, China in the end of 2019 and spread to more than 60 foreign countries as of 1 March 2020 [1]. On 20 January 2020, the first imported COVID-19 case was detected in Republic of Korea, and the epidemic curve appeared steadily until 15 February. In the second half of February, the number of reported cases increased rapidly with more than 1200 cases a week. As of 5 March 2020, there were 6088 cases confirmed including 41 deaths [1]. In Italy, the first case was reported on 6 February 2020 and the epidemic curve was steadily by 21 February. Then the number of reported cases soared rapidly, reaching 3142 reported cases and 107 deaths until 5 March 2020 [1]. To date, there are 14768 confirmed cases in the world except China, which means that more than 60 per cent of cases outside China are from Republic of Korea and Italy [1]. In this study, we modelled the early outbreak of COVID-19 in Republic of Korea and Italy to estimate the basic reproduction number under different exponential growth starting date.

## 2. Methods

We collect time series of reported COVID-19 cases in Republic of Korea from 20 January to 1 March 2020 and cases in Italy from 5 February to 5 March 2020. Following [2,3], we assumed that number of secondary cases associated with a primary COVID-19 case follows a negative binomial (NB) distribution, with means *R*_0_ and dispersion parameter *k* [3]. Here, the *R*_0_ is the basic reproduction number of COVID-19. The *k* measures the likelihood of occurrence of super-spreading events (or other factors) which could vary the growth rate. If *k* is smaller than 1, these data indicate occurrence of super-spreading events (larger proportion of ‘super-spreaders’ or ‘dead-ends’ in infected individuals) or some other factors which could vary the growth rate; otherwise if *k* is larger than 1, these data do not indicate occurrence of super-spreading events [4].

The onset date of each secondary case is the summation of the onset date of the primary case (*t*) plus the serial interval (SI). In this work, the SI was assumed as a Gamma distribution with a 4.5-day mean and a 3.1-day standard deviation (SD) [5-8]. The transmission process was simulated stochastically.

Since the number of early reported cases are stable, which appears no sign of the outbreak in January 2020, we consider the sustaining exponential growth might start since February 2020. Hence, we simulated the exponential growth starting on

- 31 January and 5 February 2020 for Republic of Korea
- 5 February and 10 February 2020 for Italy

with one seed infection. The *R*_0_ and *k* were estimated by the maximum likelihood estimates approach that fit the reported cases with Poisson-distributed likelihood framework as follows.

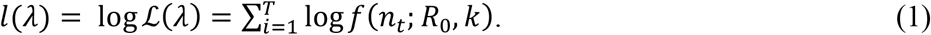

Here, the *l*(·) is the overall log-likelihood and *T* is the total number of days since the start of exponential growth. The *n*_*t*_ represents number of cases reported on *t*-th day. We calculated 95% confidence intervals (95%CI) by using the profile likelihood estimation approach determined by a Chi-square quantile.

## 3. Results and discussion

In Table 1, we estimated the *R*_0_ in Republic of Korea at 2.6 (95% CI: 2.3−2.9) and 3.2 (95% CI: 2.9−3.5) with the transmission starting date on 31 January and 5 February 2020, and the *R*_0_ in Italy at 2.6 (95% CI: 2.3-2.9) and 3.3 (95% CI: 3.0-3.6), with the transmission starting date on 5 February and 10 February 2020. Estimates of dispersion term (k) for Republic of Korea were 10 (95%CI: 5-56) and 22 (95%CI: 8-61), under the assumption that the exponential growth starting on 31 January and 5 February 2020, and for Italy were 13 (95% CI: 5-61) and 37 (95% CI: 13-61), under the assumption that the exponential growth starting on 5 February and 10 February 2020, all of which were consistent with [3] and suggested the unlikelihood of superspreading events.

**Table 1.**
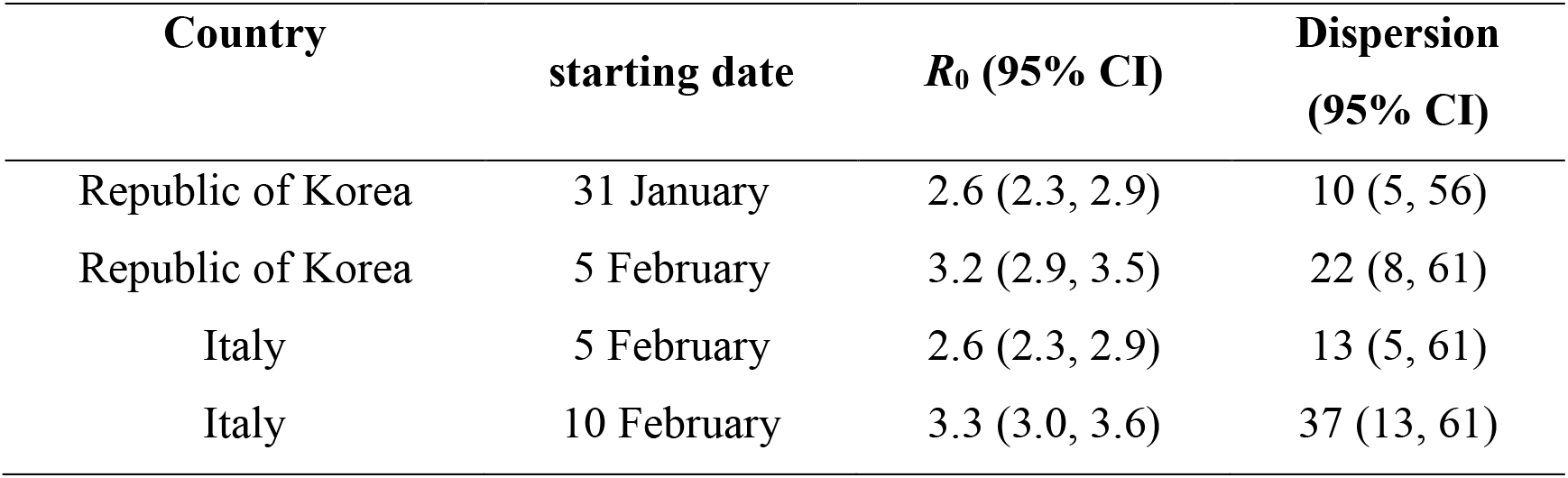
The summary of the basic reproduction number and dispersion parameter estimates under different exponential growth starting date.

| Country | starting date | $R_0$ (95% CI) | Dispersion (95% CI) |
| --- | --- | --- | --- |
| Republic of Korea | 31 January | 2.6 (2.3, 2.9) | 10 (5, 56) |
| Republic of Korea | 5 February | 3.2 (2.9, 3.5) | 22 (8, 61) |
| Italy | 5 February | 2.6 (2.3, 2.9) | 13 (5, 61) |
| Italy | 10 February | 3.3 (3.0, 3.6) | 37 (13, 61) |

Fig 1 showed that the fitting results matched the observed number of cases well. Our estimated *R*_0_ in Republic of Korea is relatively higher than that in Shim *et al*.’s [9], which set the exponential growth starting date on 20 January 2020 and the onset date data were considered. Here we use laboratory confirmation date data which covered both symptomatic and asymptomatic cases. In addition, the *R*_0_ estimates from two countries were largely consistent with those estimates based on the epidemic curve in China [10,11]. If there were evidences suggesting that the exponential growth started earlier, we note that the *R*_0_ estimates would decrease. Public activities in the late date and cold weather in our study period could have speed up the transmission which explains the higher estimates.

**Figure 1.**
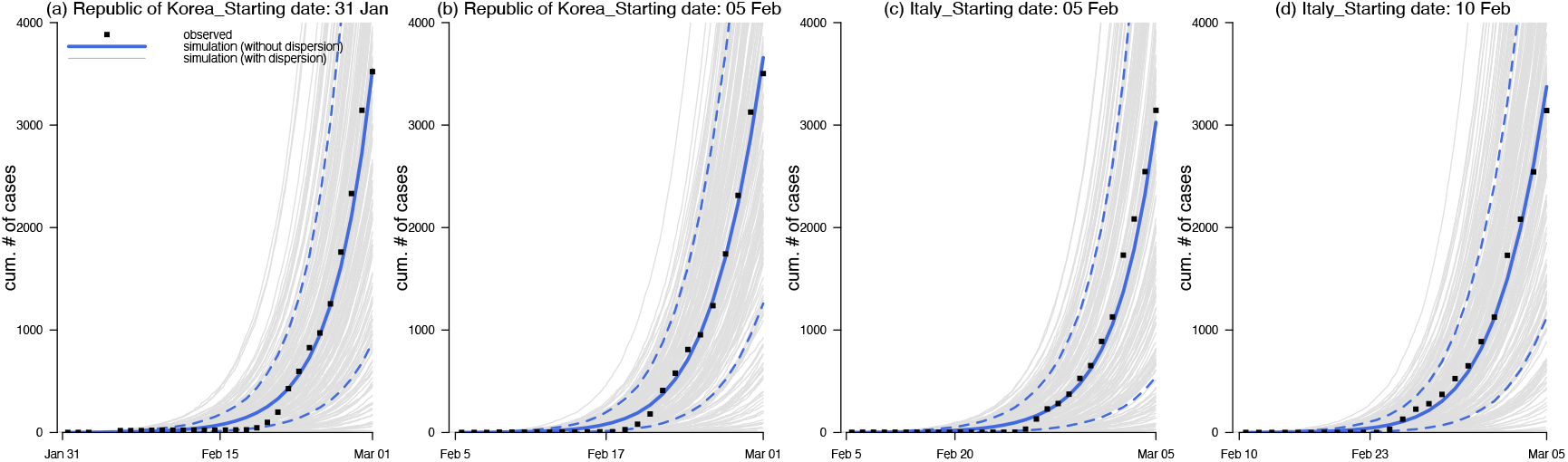
The observed (dots) and fitted (curves) number of COVID-19 cases in Republic of Korea and Italy. Panels (a) and (b) show the results in Republic of Korea with the exponential growth starting date on 31 January and 5 February 2020 respectively. Panels (c) and (d) show the results in Italy with the exponential growth starting date on 5 February and 10 February 2020 respectively. In all panels, the grey curves are 1000 simulations, the blue bold curve is the simulation median, and the blue dashed curves are the 95%CI.

The ongoing COVID-19 outbreak in Republic of Korea and Italy could be amplified due to large-scale gathering activities [12,13]. Without public health control or self-protective measures, the epidemic was likely to grow in a relatively large rate. The *R*_0_ estimates and the rapid growth of epidemic curves both indicates the disease transmissibility. Thus, control measures as well as self-administered protective actions are crucial to reduce the transmissibility of COVID-19 and thus mitigate the outbreak size and prevent for further burden. Given the superspreading is unlikely to occur, we can be confident to control the COVID-19 outbreak by reducing the reproduction number to below unity.

## Data Availability

We used only publicly available data.

## Declarations

### Ethics approval and consent to participate

The ethical approval or individual consent was not applicable.

### Availability of data and materials

All data and materials used in this work were publicly available.

### Consent for publication

Not applicable.

## Acknowledgements

None.

## Disclaimer

The funding agencies had no role in the design and conduct of the study; collection, management, analysis, and interpretation of the data; preparation, review, or approval of the manuscript; or decision to submit the manuscript for publication.

## Authors’ Contributions

All authors conceived the study, carried out the analysis, discussed the results, drafted the first manuscript, critically read and revised the manuscript, and gave final approval for publication.

